# A Software Solution for Clinical Protocol Management

**DOI:** 10.1101/2021.03.10.21253055

**Authors:** João Rafael Almeida, José Luís Oliveira

## Abstract

Clinical treatments are mostly the result of consecutive success of medical procedures. The patterns in those procedures lead to creation of clinical guidelines which are currently essential to have better health treatments. The use of electronic health record systems (EHR) helps the patient management, but it fails in the treatment guidance due to the lack of clinical decision support systems. Although these systems have decision-making features incorporated, they are not designed for treatment management and guidance. In some top edge systems, this functionality was improved and integrated, but either they are tight to the typically complex EHR. Or, many times the independent solutions lack generality to support users and disease-specific protocols.

In this paper, we purpose a decision support web tool which works independently from EHR systems. This solution allows clinicians to build and manage rule-based clinical protocols, by facilitating health care treatments, reducing time, and increasing medication accuracy. Moreover, it also enables protocol sharing among distinct institutions and physicians, creating a larger database of clinical guidelines and a network of speciliased physicians in types of diseases.

The proposed system was evaluated with the implementation of distinct clinical protocols, from different medical fields. In this validation, we describe all the steps in the transformation of a clinical guideline, from the traditional format, into a ready-to-use protocol with guidance and recommendation features.

The purposed system was developed with the collaboration of health professionals from different Portuguese healthcare institutions, which helped in the identification of system requirements and gaps of the current systems.

## 1. INTRODUCTION

Many health care treatments used today resulted, not only, from fundamental scientific research, but mostly from the continuous knowledge gathered in successful medical procedures. To simplify its understanding among professionals, these treatments usually are formalised as clinical protocols, following a set of decision rules. They are especially important during the treatment of specific diseases, such as diabetes and cancer.

The EHR systems have been designed to collect and store patient and population health information in digital format [12]. These systems have clinical data management features, such as the identification and maintenance of a patient record, medication descriptions, clinical documents, and notes, among others. However, there is a lack of decision-making and treatment-support in complex care procedures, but currently, it is recognised that EHRs require this kind of functionalities, especially in primary care units [15, 18]. The problem with this subject is the low research advances made in the past years. Additionally, the decison-making features existant have been designed for different purposes, typically used to support retrospective analyses of financial and administrative data [20, 7]. Although EHRs are advertised with decision support capabilities, this does not mean that they are designed for treatments guidance.

The use of Clinical Decision Support Systems (CDSS) for treatment management has become more eminent, and it is well-known that the use of those solutions improves the quality of patient care, disease prevention, and scientific discoveries [19]. Some disease-specific solutions have already proven their efficiency, but there is an absence of decision-making functionalities in the EHR systems. This led to the inclusion of CDSS as a required feature. Or at least include facilities to import them from external sources [21, 5].

Currently, the different approaches and solutions available, are mainly design for a specific purpose. Although some of them are designed to support multiple protocols, they have been created to integrate with specific EHR, which invalidates its diversification. This lack of flexibility is problematic when the incorporation of this type of features in health care institutions without interoperable systems is needed. The reuse of these solutions in a different context was considered a quite complex. There is also a lack of protocol sharing between institutions, which could increase the knowlegde of the physicians and be helpful in rare diseases.

This paper presents a solution aiming the clinical protocol management and treatment guidance, without being confined a specific medical field or an EHR system. The Section 2 describes what lead to the creation of this solution, as also a brief explanation of why current solutions do not fulfil the physician requirements. Section 3 describes the solution requirements and what it is expected from it. In Section 4 is presented the resulting system, explaining its architecture and the essential components. Section 5 describes the system validation, which explains how is done the protocol implementation in the system. Finally, in Section 6 the paper is concluded by discussing future work.

## 2. BACKGROUND

Clinical guidelines have been created to help health professionals during the treatment of specific pathologies. These guidelines contain general descriptions and several rules to be followed during treatments, and can be made without any computational system. However, due to the number of protocol treatments and the complexity of those treatments, the use of CDSSs was essential for management purposes.

Currently, there are some CDSS designed to perform simple but repetitive tasks, such as recognising that a laboratory test result is out of a normal range. Or that a prescribed medication should not be administrated with another that the patient is already taking [10]. The are also some specific solutions focused in a medical field, designed to guide a confined set of treatments [3, 6].

P. Fraccaro et al. [9] did a systematic review of CDSS to be adopted in multimorbidity. In this review, they identify several clinical decision support methodologies of which have interest for the purposed problem. By looking for knowledge-based systems that use rules to represent interventions from the methods chosen by these authors, only a few fit into the scope. However, these solutions have specific targets, and their purpose does not suit the problem needs.

M. Farzandipour et al. [8] also did a systematic literature review of decision support systems, that have used fuzzy logic. Exploring this review had the goal of identify one rule-based CDSS, since these authors also considered this kind of systems in their review. However, they conclude that these systems have been used to a limited extent in clinical environments.

Therefore, an attempt to identify possible solutions that fulfill the needs of our medical partners was made. Consequently, these evaluation was guided considering the following main requirements:

- User roles and activities must be controlled by an administrator, using role-based access control policies;
- The physician should be able to define protocols;
- The defined protocol most support adjustments;
- The system needs to notify health professionals about the patients monitoring schedule and treatments;
- Assigned protocols should be open for adjustments over time;
- The patient data required by the protocols should be dynamically configured over time;
- and the physician must be able to share his/her protocols with other colleagues.

### 2.1. Current methodology

The Causal ASsociational NETworks (CASNET) was the first system developed for diagnosis and treatments. This system was a prototype system created for the diagnosis and therapy of glaucoma [24]. It follows a causal reasoning approach, which is a process to identify causality, defining relationships between a cause and its effect. Currently, there is not much information available about the system.

The Open Clinical Organisation^1^ was created to encourage the use of knowledge management technologies and decision-making systems in medicine. This organisation presents some approaches to medical algorithm representation, and define formats for modelling clinical guidelines. The GLIF3 is a language designed for this propose, supporting computer-based guidelines. It establishes several characteristics in the workflow such as the logic rules, clinical data of the patient and the actions to perform in the treatment. To support this language, it was developed the GLIF3 Guideline Execution Engine (GLEE). GLIF3 allows the configuration and execution of guidelines, and keeps the patient data in the system, or in an EHR system, making the system hybrid [23]. However, it is not available any system deployment to be tested, and according to the documentation, the system is outdated.

The GESDOR follows the same approach used by GLEE, but in contrast to the GLEE, it uses generalised guideline formats [22]. Therefore, the guidelines can be encoded in different structures and stored in distinct repositories. The problem with this methodology is that it was only a theoretical model. This theory did not lead to any practical approach. Therefore there is no tool available.

The OpenCDS^2^ is an open-source framework, which provides a Java-based engine, designed as a Decision Support Service (DSS). This framework is a service-oriented system, without Graphical User Interface (GUI) [14, 16]. The absence of proper documentation to create a web client or to integrate with others systems discouraged its use.

The Immunization Calculation Engine (ICE) is an open-source system designed for the forecast immunisation, and it has two major components.The core component that evaluates the patient’s history and generates a recommendation, which is designated as ICE Web Service. This component runs using the OpenCDS middleware. The second component is the Clinical Decision Support Administration Tool (CAT), which is a GUI framework design to present the OpenCDS resources. This project is a composition of different systems, new in the market, and therefore the use of it was discouraging. Additionally, this system was designed for the forecast immunisation and to use it in another scenario, requires some efforts.

All those described systems are open-source, with useful features, but they did not fulfil our needs. Then, we decided to explore some commercial solutions. The IndiGO is clinical guideline system, which follows the Archimedes model and it has several clinical variables to store the patient’s records [4]. This system also allows the creation of individual protocols, customised for each patient, and it provides methodologies to be integrated with the EHR system. However, there is a lack of documentation, maybe due to the system license. Also, we cannot be able to test this system.

### 2.2. The GenericCDSS system

The primary focus was to use an open-source solution to support the initial requirements. Or if the system did not has all the features, at least allow the extension of new ones. However, autonomous CDSS, i. e, without the support or integration of an EHR system are a subject underexplored. There are some solutions but designed for a specific goal. Another issue found was the discontinuance of technical support in some of these solutions. They have the potential to fulfil the defined needs, but they use old technologies and do not support multi-platforms, such as mobile devices.

In the past, we developed a CDSS designed for diabetes treatment [1]. What then lead to the development of a multipurpose CDSS, which is now an open-source solution [2]. The purposed system is extension based in the GenericCDSS, with new features and goals.

## 3. METHODS

### 3.1. Requirements

In the previous section, the fundamental requisites of health professionals were described, which needs to be incorporated into the system design. Regarding these needs, the following set of functional requirements were defined to address the system architecture:

- **Managing Protocols**: Physicians should be responsible for protocol management. They can create, read and delete the protocol and its information. However, the administrator should be able to help the physicians in some situations. For instance, when the protocol has a complex structure, the physician could request for the administrator help in the implementation stage. Regarding this, we decide that physicians and system administrators could have protocol management permissions.
- **Searching Protocols**: Physicians need to be capable of searching for protocols in the system. This feature is essential when the system has a high number of protocols, or to check if other physicians have already introduced the desired protocol. It should be possible to search for the protocol using different criteria, such as creation date, name or description. Also, nurses are not allowed to search for protocols, to avoid misinterpretations.
- **Assigning Protocols to Patients**: When the patient is admitted to the hospital, or even later, the protocol assignment should only be made by the physician. Nurses do not have permissions to take this kind of decisions.
- **Customising Protocol**: Despite protocol edition be associated with the protocol management, this action has extra importance. Protocol customisation focus on some adjustments made in the structure of an assigned protocol. This changes cannot interfere with the remaining assignments of the same protocol to other patients. For the system, it is a fork of the original protocol. Additionally, only physicians can perform such customisation.
- **Seeing Protocol Notifications**: The system needs to notify the users about the protocol execution state. This requirement is essential for patients follow-up, namely made by the nurses. This feature increases the measurements accuracy.
- **Seeing Assigned Protocols**: A dashboard with all the assigned protocols to each patient is also required. This information should be available to physicians and nurses. It helps nurses to know what is the patient state quickly, and if the treatment was already done. Also, it aids the physicians to understand what has been done by the nurses. This track improves the identification of protocols adjustments, when necessary.
- **Patient Clinical Data**: There are two different stages about the patient clinical data: 1) the clinical variable definition, discussed later in this section; and 2) the patient data insertion, the moment that the data is added in the system by both entities. Some of this information is given by the physician when s/he checks the patient state. However, nurses also need to introduce patient data during the treatments.
- **Managing Patients**: The physicians are responsible for performing the patient admission and dismission from the system. In this process, the patient data is inserted or updated into the system. Nurses can see the patient information, but they are not allowed to discharge the patients, either to admit them in the hospital.
- **Patient History**: All the patient data, protocols assignments and execution results must be kept in the system. Physicians and nurses have read permissions to access any patient information. However, changes in this history are not allowed by any entity.
- **System History**: This feature is designed for the system administrator. S/he is the only one that needs to be able to navigate through the system’s history, which will allow him/her to see if the system is stable and functional.
- **Managing Clinical Variables**: After the system’s installation, the patient’s clinical variables must be defined. Since it is back office activity, and to assure the system’s stability, this task should only be performed by administrators. However, if a physician needs more clinical variables, s/he should request it to the system’s administrator.
- **Managing Users**: All users in the system, physicians and nurses, are managed by a registered administrator. Therefore, nurses and physicians do not control other users activities in any situation.

### 3.2. System Description

The system propose in this paper was designed concerning different workspaces to cope with the distinct users roles. In the profile nurse, the first view is a dashboard with a list of patients admitted to the hospital, with some essential information about each patient. In this view, the nurse can execute the assigned protocol, or only verify the patient state.

The protocol execution presents the recommended prescription for the patient, considering his/her current clinical state. Then, this data can be later explored in the patient view. This procedure is the system goal, which will automatically consult the clinical guidelines to identify the right dosage or perform the calculations that before have been done manually.

The physician can also access all these dashboards, and execute the protocol if s/he desires. Also, there is a dashboard with the protocols in the system, and a protocol editor, design to create or updated these clinical protocols.

The administrator has a different workspace, with new dashboards where it is possible to manage all the system. In these workspaces, s/he can maintain the clinical variables, user roles, and other modules belonging to the system.

## 4. RESULTS

### 4.1. System architecture

The system follows a Client-Server model, in which each side is subdivided in several layers (Figure 1). This figure presents the system’s architecture, where this decoupled approach is emphasised, namely the core modules and the extensions elements, without having to interact with the rest of the system.

**Figure 1.**
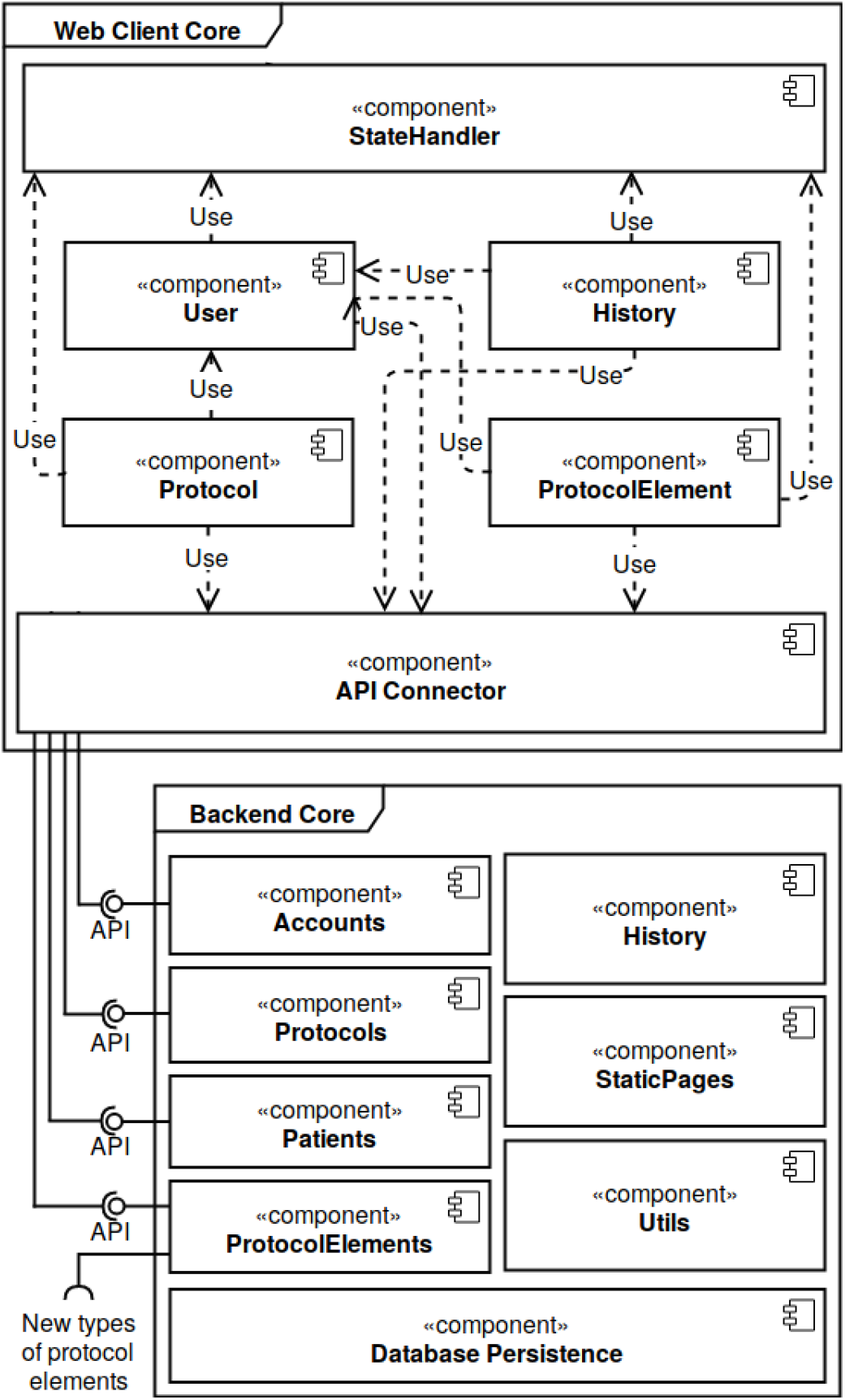
The system architecture, which follows a client-server model. The web client core has several components to handle with the visual interface and the communication with the backend. The backend core is composed of several components and some APIs to allow external accesses.

The Client-side encapsulate the presentation element of the system, divided into: 1) presentation layer, which is responsible for the user interfaces; and 2) controller layer, that allows the interface to access data, by consuming the backend web services (API Connector).

The Server, which is mainly the backend core, is subdivided into three sub-layers: 1) business; 2) persistence; and 3) service provider. The persistence layer is responsible for storing and maintaining the system’s data. The business layer contains most of the application’s logic. Finally, the service layer provides a RESTfull API with services prepared to interact with all the system’s functions with or without the client. This layer will be used by the client to access all the core features.

#### 4.1.1. Client side

The client was built keeping the modularity and extensibility in mind. To archive this, we combined ReactJS with RefluxJS technologies as the base of the client interface. This combo has a unidirectional data flow, which leads to a well-known pattern composed of actions and data stores. Figure 2 represents this logic, where actions launch new data, which will cross the stores before being rendered in the component view. The stores are responsible for handling the data changes, and to perform the backend calls. The views send signals using the actions available to fetch the data. This flow simplifies and isolates the communications with the backend, without incorporate that logic with the component’s core.

**Figure 2.**
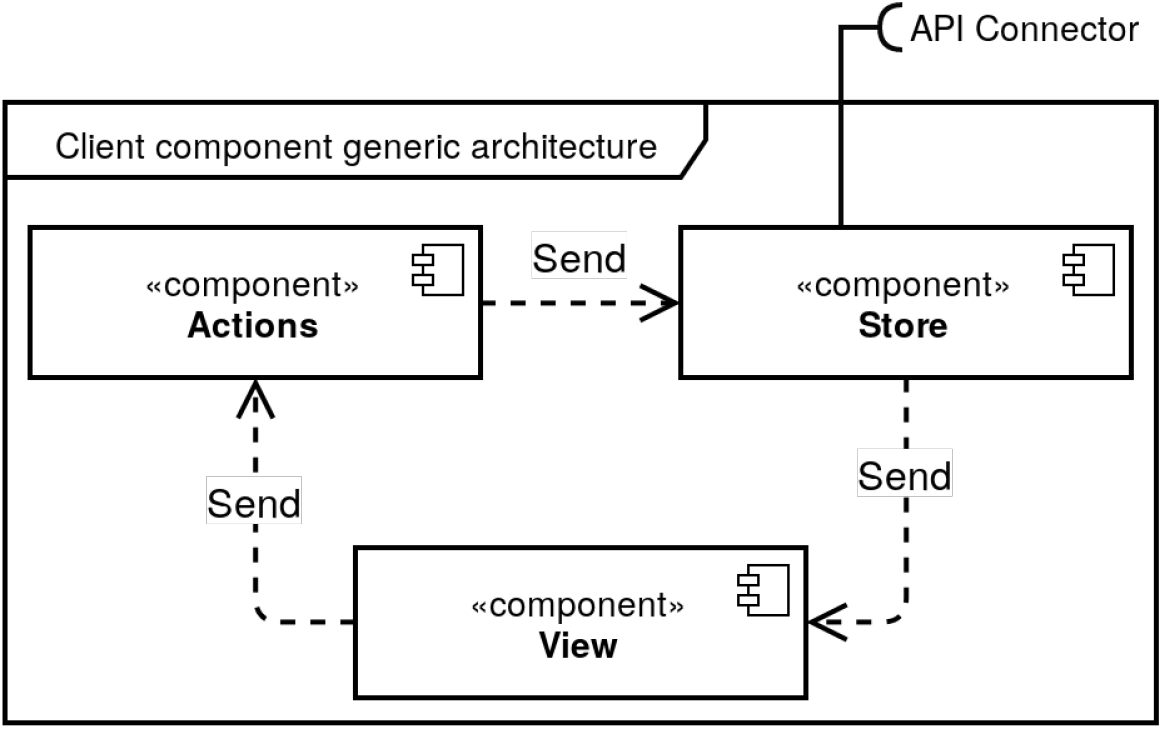
Client component architecture. It is represented the client flow of interactions to get and display the data in the user interface

The communication with the backend made in the stores is performed using the web services. These web services are called by the API connector (Figure 1 - Web client core), responsible for making the connection between the client and the server, with the support of Axios^3^. This framework is an HTTP client for browsers, and the communication between both sides follow the steps below:

1. The web component needs some data stored in the server;
2. This component sends an action signal defined in the module responsible for providing the desired data;
3. The action triggers the data store.
4. The data store request the API Connector to consume the web service, providing the necessary information about it;
5. The API Connector communicates with the server side and receives the response;
6. The data store receives the data and updates its state;
7. Finally, when the component state changes, the view fetch the requested data.

#### 4.1.2. Server side

Similar to the client side, we also kept the modular architecture in the backend core. This core was reduced to the minimum but allows its extensibility through new plugins.

In Figure 1, in the Backend core section, it is represented all the backend components. This architecture was developed using Django^4^, which is an easy-to-use programming framework to build web or mobile applications [17]. This framework is recognised by its simplicity, allowing the creation of components without unnecessary complexity [11]. The main reason for choosing this framework is the excellent documentation and the support that it provides.

Another Django characteristic is the isolated and decoupled components, which significantly simplifies the integration of a micro-kernel architecture. Additionally, the framework uses PIP, a package manager to keep track of which packages are installed. This simplifies the integration with third-party components. Since Django has a big open-source community of contributors, our system uses several modules adopted from outside developers.

### 4.2. Patient Data

Despite the system being design for protocol management, it needs to manage the patients as well. The information base to execute the protocols is the patient state. In other words, the patient clinical variables will determine the protocol behaviour. However, we wanted a system generic and decoupled from any EHR system. This requires a minimum of features to manage the patient information.

Therefore, we design a module to manage this kind of data. In this module, we handle with some patient information, such his/her name, birth date and contact. But we also define the clinical variables need in the future to supply the protocols. These variables must allow changes, without the need of recompile all the system, or change the database structure. Concerning this, we create an architecture that permits the configuration of patient clinical variables in run time. These variables are organised in groups, and the administrator makes all these configurations.

Currently the system offers three different types of variables: 1) numeric type, where it can be inserted real numbers; 2) text variables, which accept text and numeric values, but for numeric use, it is strongly recommended the numeric type; and 3) optional type, that has a set of options, also defined by the administrator. The users can choose one of these. This architecture gives to the system all the necessary features to manage the patient data, that will sustain the clinical treatments.

The Figure 3 represents the individual patient view. In this figure we can see: a) the personal patient information, namely the patient name and contact; b) the different patient information; and c) the group of clinical variables. The patient information represented in b) are the clinical variables, the performed treatments, which is a history of all the interactions made with the patient, and the assigned protocols. The groups of clinical variables represented in c) are the clinical variables discussed before with some measurements. This section also has filtering and sorting features.

**Figure 3.**
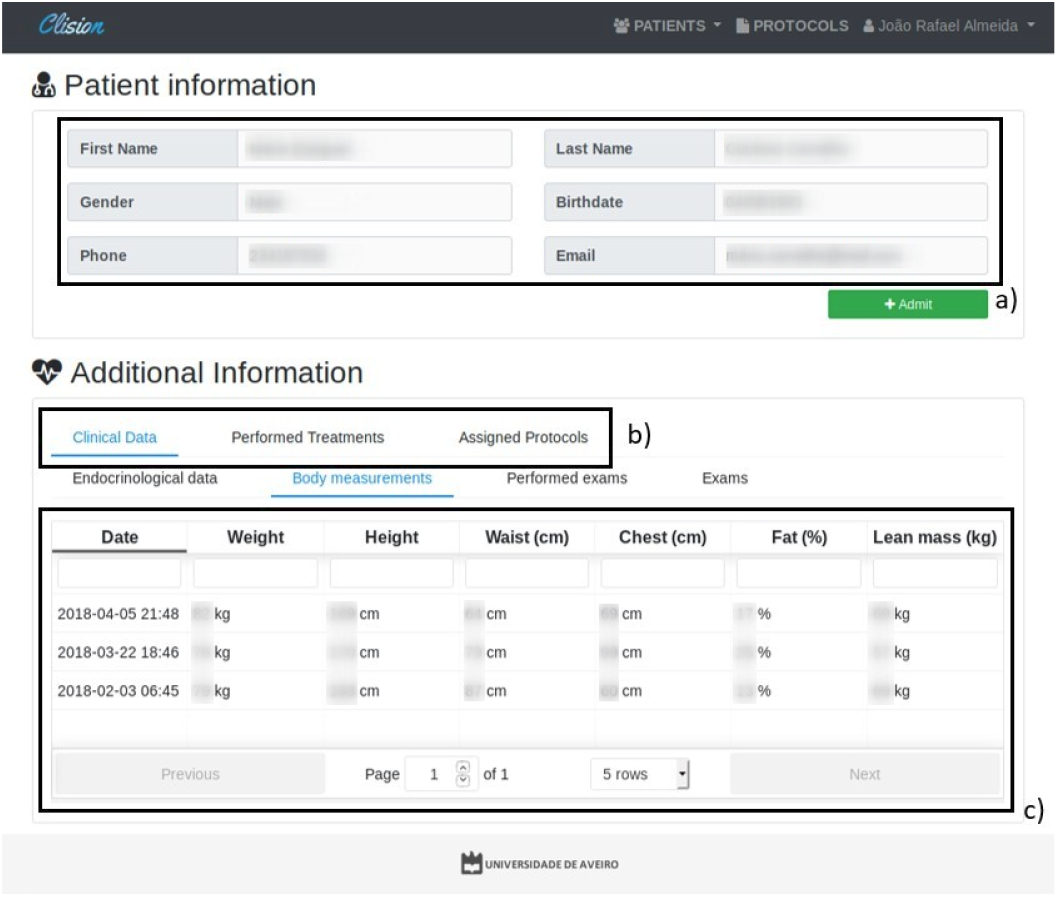
Individual patient view. This figure represents: a) the personal patient information; b) different patient information, such as the clinical variables, the performed treatments, and the assigned protocols; and c) the clinical variables.

### 4.3. Protocol Structure

After the clinical variable definition, the system can now accept the creation of the clinical protocols. Architecturally this requires two different modules: 1) the protocol module, which has the main protocol information, such description and schedules; and 2) the protocol elements module, that has the different types of elements which compose the protocol structure. The protocol module also handles with the protocol execution/assignment.

Currently, the system has three distinct protocol elements, which are: 1) Inquiry elements, with the function of collecting patient data (clinical variable insertion); 2) Decision elements, that compares the inserted data in the inquiry elements with conditions, influencing the protocol behaviour; and 3) Action elements, which provide the treatment recommendation. A protocol is built following the composition of several of these three elements in a flow structure. These structured elements create a protocol template, which is then assigned to patients and executed numerous times. Furthermore, the system currently only have three protocol element types, but it allows the inclusion of more elements. For instance, including hardware integration to simplify the recorded patients clinical data using medical devices.

The protocol module manages all the protocol assignments and schedules. This module keeps all the execution steps for each protocol assignment. Also, it has a mechanism to efficiently represent the schedule option in a language understandable by the health care professionals. These schedules are shows as time-labels defined by the administrator, with the moment of the day that the protocol should be executed. The main reason for this is because protocols have medical terms related to the non-time factors, such as meals or bedtime.

The Figure 4 represents the system view for the protocol editor. This figure shows the Hypoglycemic protocol implementation, and it highlights two essential system features: a) the protocol schedule, following the time-label definition; and b) the protocol definition, that describes the protocol elements, following a workflow structure.

**Figure 4.**
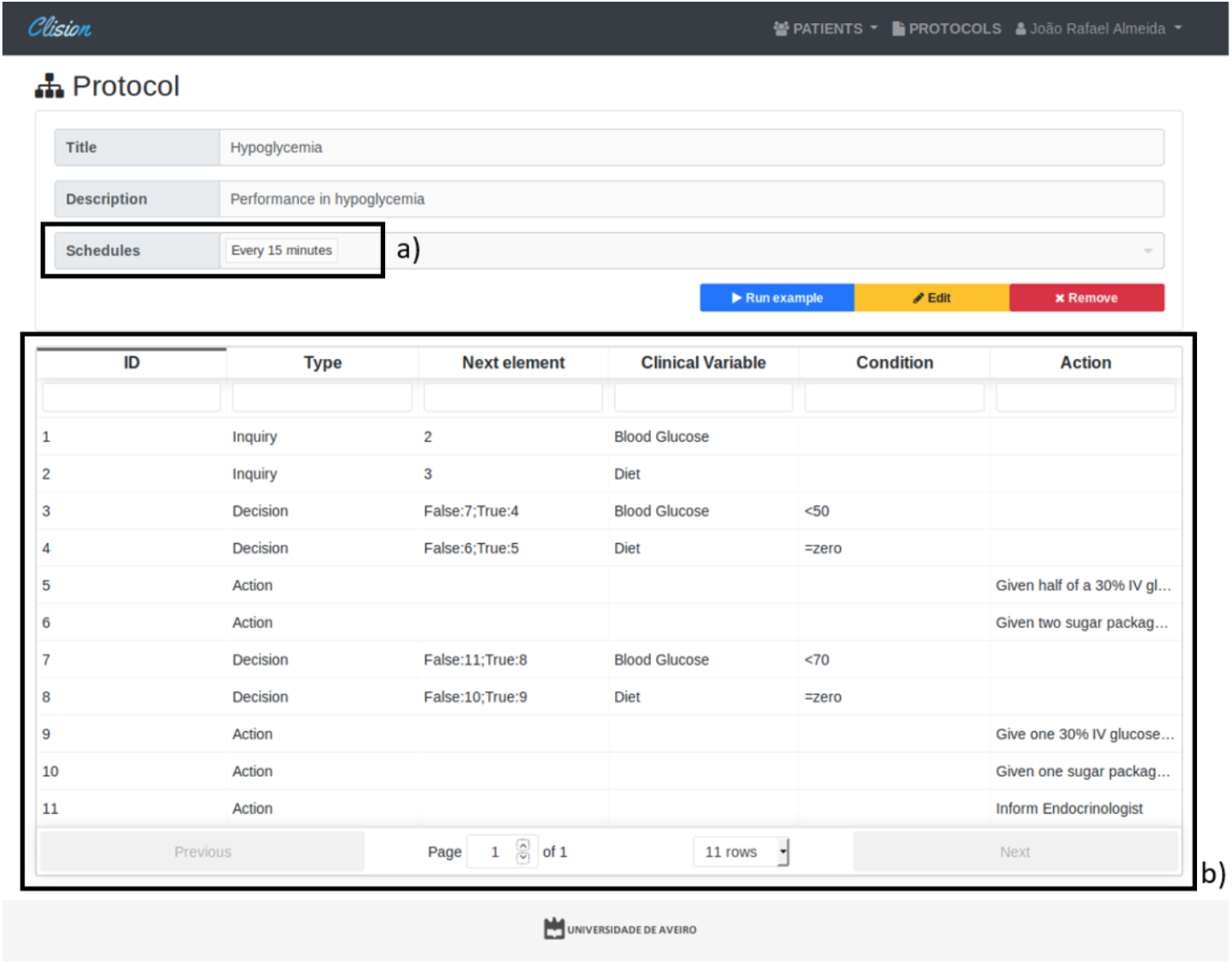
The Protocol editor view. This figure represents: a) the protocol schedule, which follows a time-labels definition; and b) the protocol definition, that describes the protocol elements.

### 4.4. CRUD Operations

The acronym CRUD further the Create, Read, Update and Delete operations on the database. These operations are usually the base actions of a relational database in the persistence layer. However, in our system, some of these operations have been disabled for the users to avoid losing valuable data.

The typical users of the system are nurses and physicians. These roles do not have delete permissions to any components in the system. The remove function in some views only hides the data, but all the history is kept. Only administrators have this privilege.

The patient clinical data is created only when a protocol is executed, which means to insert new entries of patient data, a protocol needs to be assigned and executed. The create operation for this module is restricted to this condition, which forces the patient admission in the system.

These restrictions in the CRUD operations create a pipeline to ensure the system stability and the data consistency. Also, all these operations are made through web services, which will be described below.

### 4.5. RESTful API

The system API conforms to the REST architectural style. We used Django REST Framework^5^, which is the recommended solution to build RESTfull API over Django. This framework has detailed and well-written documentation, with excellent community support, similar to Django. It also supports authentication policies including the packages OAuth1a and OAuth2, and serialisation mechanisms.

**Figure 5.**
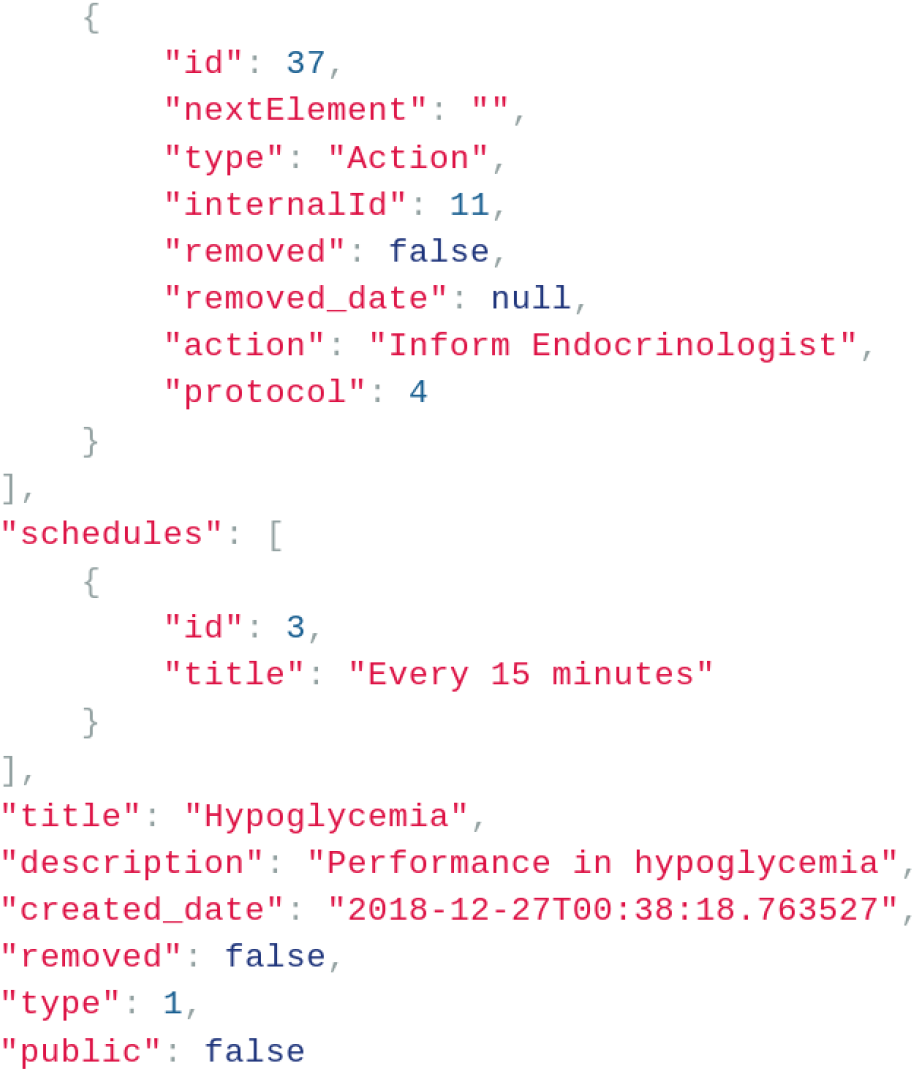
Part of the RESTFull web service response for a GET request to a clinical protocol.

Each module of our system has its data models and views. These views have serializers to handle with complex data such as query sets and model instances, which can then be converted to native Python data types. Also, each view has defined filters with several criteria, and data sorting to be used by the client side. The web services responses follow the format of key-value pairs, based on JSON. The Figure 5 represents part of the GET response for the protocol presented before in the protocol editor.

These views have an URL defined following some rules. These rules aim the homogenization of the web services definition in all the system modules. This methodology helps developers to quickly understand from which module, model, and view, the method has been called. Therefore, all web service starts with the path *‘/api*, followed by the module’s name and view. Then, the next element in the path can be the method to call or the id, depending on the web service’s definition.

The listing web services accept parameters in the request to characterise the response. We can define the page number to be retrieved or the number of records on if each page. The definition of those parameters adopted the following format: 1) ‘?page=<x>‘ for page number; and 2) ‘?page size=<x>‘ for page size. Also, for each listing web service it is incorporated filter and sort features, through the use of ‘?<field>=<value>‘ and ‘?ordering=<field>‘, respectively.

### 4.6. Protocol Sharing

Converting clinical guidelines to digital format has more advantages than optimise the treatments, with this system, is also possible to easily share the protocols between different institutions. The protocol template can have unrestricted access since it does not contain any sensible patient data, only the protocol structure, which are already public in the literature. Therefore, when a physician defines a protocol template, with all the steps and clinical dosages, s/he can quickly share it.

This feature can improve the protocol methodology by having a group of people specialised in one pathology, analysing the protocol. This way, we have a community distributed in several healthcare institutions, with different resources and backgrounds, capable of contributing to the protocol composition.

## 5. DISCUSSION

In this section, we explore several challenges to test and validate the feasibility of our solution as an autonomous clinical decision support system. The purposed solution aims the clinical protocols management, and to validate it, we present an use case originated from the needs of different health care institutions.

### 5.1. Case Study

Some Portuguese healthcare institutions have EHR systems installed with decision-making features, but not designed for treatment guidance. Our case study is based on this need, namely systems without clinical protocol management and treatment electronic guidance features. To accomplish this goal, without developing a custom solution for each institution, we create this system.

Some distinct protocols, from different medical fields, have been used to validate the solution. To assess the system, we defined four key goals:

- Protocol conversion to digital format;
- Treatment guidance;
- Automatic therapeutic recommendation;
- and patient and treatment history for future references.

### 5.2. Validation

In the validation of all the features, we deployed a prototype in a controlled environment. Then, we insert some protocols currently in use in several medical units of different diseases, namely:

1. Hypoglycemia treatment;
2. Contrast-induced nephropathy treatment;
3. Preoperative;
4. Anticoagulant therapy;
5. and Postoperative pain relief.

The protocol insertion in the system requires that text guidelines must be interpreted and defined into a flow structure. To explain the protocol setup, we will consider the Hypoglycemia protocol, which is also available in our demo installation. This protocol has a small set of conditions that we detail belong. Then, we analysed the protocol and described what is necessary to convert it into a more friendly format.

#### 5.2.1. Hypoglycemia

The Hypoglycemia treatment is a fast-acting protocol, and the measurement’s periodisations are short, i.e., measures should be made every 15 minutes if the blood glucose patient is less than 80 mg/dl.

The protocol methodology only requires the knowledge of the patient’s diet and his/her blood glucose values. When the patient can eat, s/he receives one sugar packets (which is about 6 grams of sugar), only if the blood glucose value is between 50 and 70 mg/dl. However, if this value is less than 50 mg/dl, the patient receives two sugar packets. A patient on a zero diet receives half of a 30% intravenous glucose ampoule and glucose serum 5%, when the blood glucose value is between 50 and 70 mg/dl, and a 30% intravenous glucose ampoule and glucose serum 5% when it is less than 50 mg/dl.

From a medical point of view, the protocol description lies in what has previously described. To deploy the protocol in the system we need to define some clinical variables, which supports its execution. These variables are:

- **Blood Glucose:** This clinical variable is the numeric type. In this protocol, it influences the protocol execution. When it’s value is less than 80 mg/dl, the application of this protocol is required.
- **Diet:** This clinical variable is option type, where we define if the diet is: 1) normal; or 2) zero, if the patient is not eating.

The description above can be represented in a flow chart. Figure 6 represents all the steps and possible behaviours for the protocol. The user that executes this protocol does not have the perception of its complexity since all these steps are hidden. S/he only introduces the inquired data and receives a set of instructions.

**Figure 6.**
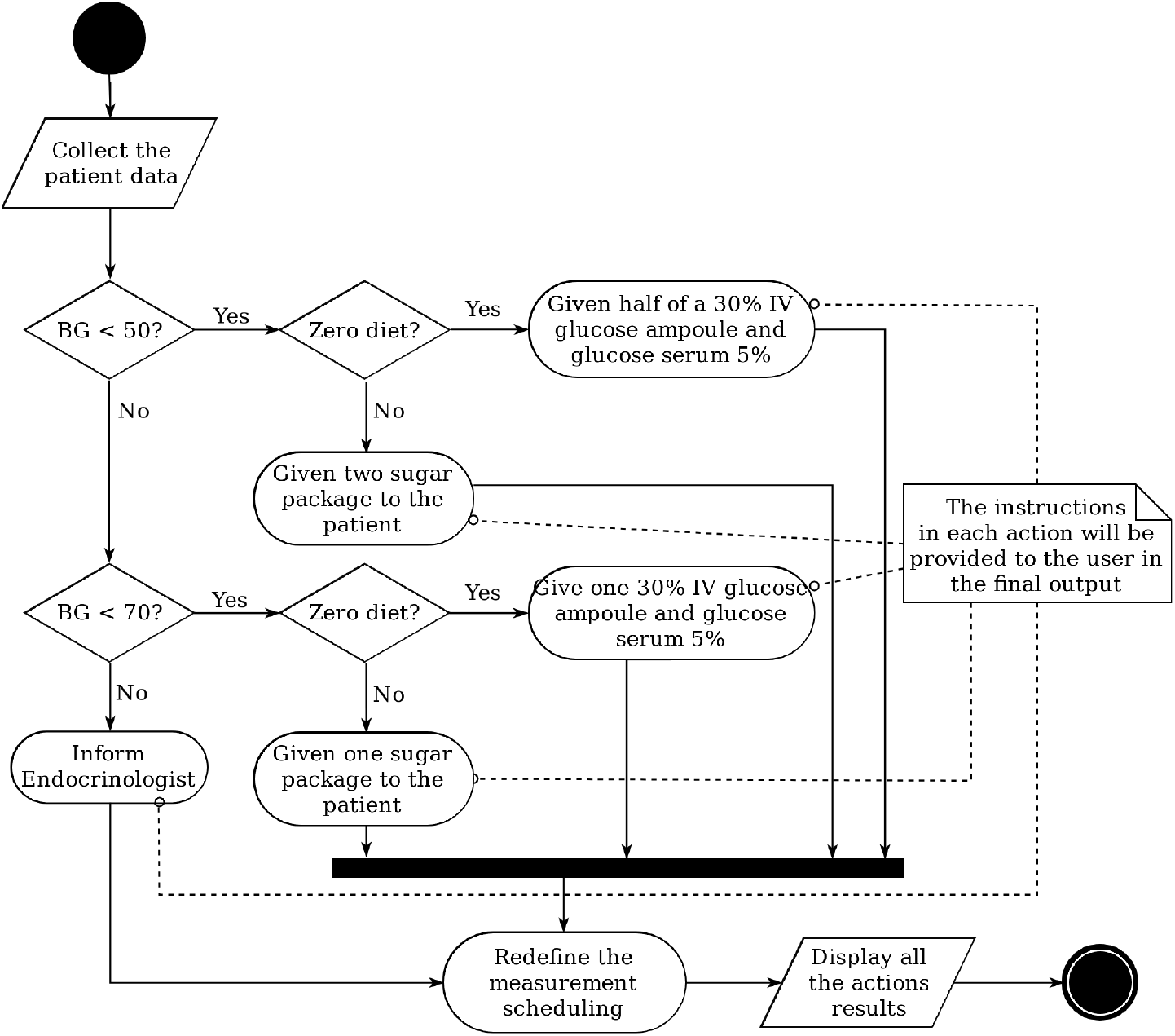
Flowchart of the hypoglycemia protocol.

Therefore, for each protocol we define a similar flow chart to easily insert the it in the system. The hypoglycemic protocol described before, is represented in the system in the Figure 4.

Despite being a preliminary assessment, the system can support clinical guidelines, if they are properly converted to an identical structure of the described protocol. The effort in the setup phase will return benefits because the treatments will be more accurate, will have a history, and the time spent interpreting the text guideline is abolished

## 6. CONCLUSIONS

Clinical guidelines are fundamental in hospital institutions to support the treatments. These guidelines ensure the quality control of the medical procedures. However, medical research is growing, and new therapies are always found. This evolution creates needs, such a flexible ICT to incorporate these decision-making guidelines.

Currently, the new EHR already have CDSS incorporated, but old systems without good decision-making capabilities are impotent to this growth in the medical field. The options that they have are: 1) improving all the systems in the institution, which has high costs; or 2) use the traditional paper to manage all the guidelines and treatments.

Our proposal is designed for these institutions, with this gap in their ICT features. The main scientific contribution of this paper is the system itself that hopefully will increase the healthcare quality, and the clinical guidelines sharing feature. This idea of community could lead to new and better protocol treatments. Currently, we only have the protocol sharing features. However, there is space to enhance the system for patient data sharing, allowing the observational studies, without infringing their privacy [13]. This feature could help in the protocol efficiency analysis, but this subject will not be discussed in this paper.

Therefore, we plan to make an installation in the partner institutions that collaborate with us in this work. The goal is to interact with real patients, replacing the traditional use of paper. A demo version of this system, with all the permissions, is available at https://bioinformatics.ua.pt/clision.

## Data Availability

No datasets were used in this manuscript

## Footnotes

1 http://www.openclinical.org/

2 http://www.opencds.org/

3 https://github.com/axios/axios

4 https://www.djangoproject.com/

5 http://www.django-rest-framework.org/

